# Biological attributes of age and gender variations in Indian COVID-19 cases: A retrospective data analysis

**DOI:** 10.1101/2021.02.13.21251681

**Authors:** Savitesh Kushwaha, Poonam Khanna, Vineeth Rajagopal, Tanvi Kiran

## Abstract

**Background:** The associated risk factors, co-morbid conditions and biological variations varying with gender and age might be the cause of higher COVID-19 infection and deaths among males and older persons. The objective of this study was to predict and specify the biological attributes of variation in age and gender-based on COVID-19 status (deceased/recovered).

**Methods:** In this retrospective study, the data was extracted from a recognised web-based portal. A total of 112,860 patients’ record was filtered out and an additional 9,131 records were separately analyzed to examine age and gender relationship with patient’s COVID-19 status (recovered/deceased). Chi-square, t-test, binary logistic regression, and longitudinal regression analysis were conducted.

**Results:** The male COVID-19 cases (65.39%) were more than females (34.61%) and mean age of infected and recovered patients was 39.47±17.59 years and 36.85±18.51 years respectively. The odds for infection was significantly higher among females for lower age categories, which declines with age. The age-adjusted odds for recovery were significantly higher among females (O.R.=1.779) and odds for recovery was highest in 5-17 years age category (O.R.=88.286) independent of gender.

**Conclusion:** The chances of being COVID-19 infected was higher for females of lower age categories (<35 years) which decreases with age. The odds for recovery among females was significantly higher than males. The chances of recovery declines with increasing age and the variation could be attributed to the biological differences between age categories and gender.

## Introduction

The COVID-19 pandemic has challenged the global health systems and medical sciences. The SARS-CoV-2 (Severe Acute Respiratory Syndrome Coronavirus-2) virus has infected about 10 per cent of the global population and since the end of May 2020, India is facing an upsurge of COVID-19 cases and has been ranked as 2^nd^ most affected country in the world (1). Several assumptions were made for the differential impact of COVID-19 on age and gender. The environmental and socio-economic factors was also utlised for depicting the vulnerability to COVID-19 (2, 3). The sociological factors like jobs, daily activities, economy, literacy etc. put individual differentially towards diseases (4, 5). The susceptibility to external pathogen also differs due to biological differences at various age (6) and between the genders (male and female) (7).

The immune system plays a crucial role in the prevention from various microorganisms, including viruses. The normal human immune system adapts during the fetal to infant stage, matures during adolescent to the adult stage with variability during pregnancy and decreases as the senescence approaches (8). These fluctuations in the immune system throughout life poses a higher risk for complications in infants, pregnant women, and elderly. There are several components involved in differentiating of immune system based on gender and age. The variation in the level/count of immunoglobins, CD4 and CD8 cells, B-cells, T-cells among males and females might be causing the variation in COVID-19 cases and deaths (6, 7, 9-12).

The publication of studies stating the binding of SARS-CoV-2 with Angiotensin-converting enzyme 2 (ACE2) (13) and some studies claiming a strong binding affinity to human ACE2 (14), evoked a debate on continuation of ACEIs and ARBs (Angiotensin II receptor blockers) drugs among Cardiovascular Diseases patients. However, there is a lack of evidence to prove a significant increase in ACE2 levels, subsequent ACEIs and ARB’s dosages (15). Due to COVID-19 the association of ACE2 with gender and age also came in light after some studies showed differences in COVID-19 infection and death rates based on age and gender (16).

The COVID-19 positive cases are continuously increasing in India, and little evidence is available highlighting the age and gender perspectives of this disease. Therefore, the present study was an attempt to study the association between age gender and patients’ status (recovered/deceased) among positive COVID-19 cases and also discussed the possible biological reasons for the variation among different age categories and gender. The present analysis is expected to provide evidence for framing age and gender-specific public health policies and treatment of COVID-19 infections.

## Methods

The present study is based on secondary data which was extracted from a recognised web-based portal (https://api.covid19india.org/documentation/csv/), which is the open-source portal/page of COVID-19 tracking website (https://www.covid19india.org/). The portal keeps a real-time track record of COVID-19 cases in India and allows the viewer to access the raw (MS Excel) form of data under various categories like time-series cases, state-wise, district-wise etc. The data available on the portal was in accordance with the official estimates and was sourced mainly from various government websites, government Twitter accounts updates and newspapers. The time-series cases data available in MS Excel Sheets from ‘raw_data1’ to ‘raw_data16’ were downloaded. The cases reported between 01-03-2020 to 30-09-2020 (7 months) were filtered in a separate file. The raw and coded data was uploaded to Mendeley data (17). A total of 112,860 records were separated based on nationality, date of COVID-19 positive, age, gender (male/female), and patient’s COVID-19 status (recovered/hospitalised/deceased). From these extracted records, 9,131 records were separated based on the status (recovered/deceased) of COVID-19 patients.

## Data analysis

A retrospective secondary data analysis was performed for COVID-19 cases in India. Descriptive statistics were used for reporting the mean, standard deviation, percentage and count under various categories and sub-categories. After cleaning and coding of data, appropriate statistical analysis was performed for group comparisons and estimating relationships. The age and gender records for N=112,860 and age, gender, and patient’s status records for n=9,131 were analysed separately (Figure 1). The age was divided into five categories (0 to 4 years, 5 to17 years, 18 to 35 years, 36 to 55 years and 56 years & above). The recovery and deceased estimates were not in accordance with national estimates therefore, longitudinal regression modelling was conducted for n=9,131 records. The chi-square test was used to estimate the differences between categorical variables (age categories, gender) and independent t-test was performed to estimate the differences in mean values of the continuous variable (age) across gender. The binary logistic regression analysis was also used to examine the relationship between the dependent and independent variables. The Odds ratio (ORs) along with 95% confidence interval was used to assess the risk among gender and age.

**Figure 1.**
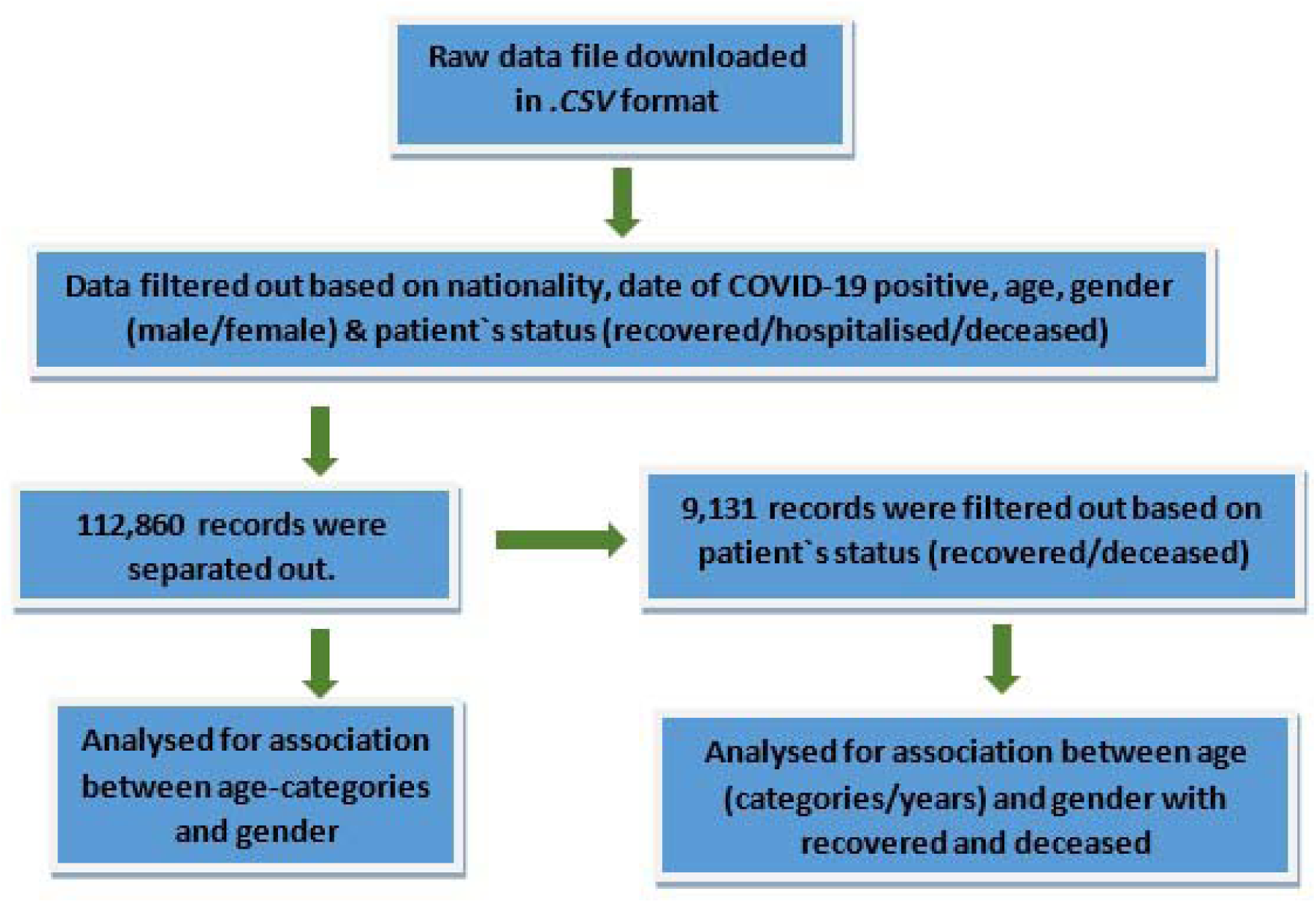
STROBE flowchart of study methodology.

For all these analyses the Females and Males were coded as ‘1’ and ‘2’ respectively. The deceased was coded as ‘1’ and recovered as ‘2’. Among age categories, the lowest age category i.e., 0-4 year were coded as ‘1’ then, 5 to 17 years as ‘2’, 18 to 35 years as ‘3’, 36 to 55 years as ‘4’ and 56 years and above as ‘5’. The statistical significance was considered at *p-*value of <0.05 and all the statistical tests were performed using IBM SPSS Version 26.0.

## Results

The extracted data includes the details of 73,797 (65.39%) males and 39,063 (34.61%) females COVID-19 patients. Most COVID-19 cases (37.48%) were found in the 18-35 years age category (Table 1). The Highest distribution of infected male (37.21%) and female (37.99%) patients was within 18 to 35 years age category and the lowest distribution of male (1.28%) and female (2.19%) patients were found among 0 to 4 years age category (Figure 2). Moreover, the difference between gender among age categories was found to be statistically significant (p<0.001) (Table 1). The mean age of all COVID-19 patients was 39.47±17.59 years and the difference between the mean age of males (39.98±17.19 years) and females (38.50±18.29 years) was also statistically significant (p<0.001) (Table 1), with a mean age of COVID-19 infected males being significantly higher than that of the female counterparts.

**Table 1.** Showing mean age and age-categories wise comparison of male and female COVID-19 patients.

| Age categories/Gender | Male | Female | Total | p-value |
| --- | --- | --- | --- | --- |
|  | Mean±S.D./<br>N (%) | Mean±S.D./<br>N (%) | Mean±S.D./<br>N (%) |  |
| Age | 39.98±17.19 | 38.50±18.29 | 39.47±17.59 | <0.001 |
| 0-4 years | 946 (52.53) | 855 (47.47) | 1801 (1.60) | <0.001 |
| 5-17 years | 4457 (56.98) | 3365 (43.02) | 7822 (6.93) |  |
| 18-35 years | 27459 (64.91) | 14841 (35.09) | 42300 (37.48) |  |
| 36-55 years | 26545 (68.07) | 12454 (31.93) | 38999 (34.56) |  |
| 56 years & above | 14390 (65.59) | 7548 (34.41) | 21938 (19.44) |  |
| Total | 73797 (65.39) | 39063 (34.61) | 112860 (100.00) |  |

**Figure 2.**
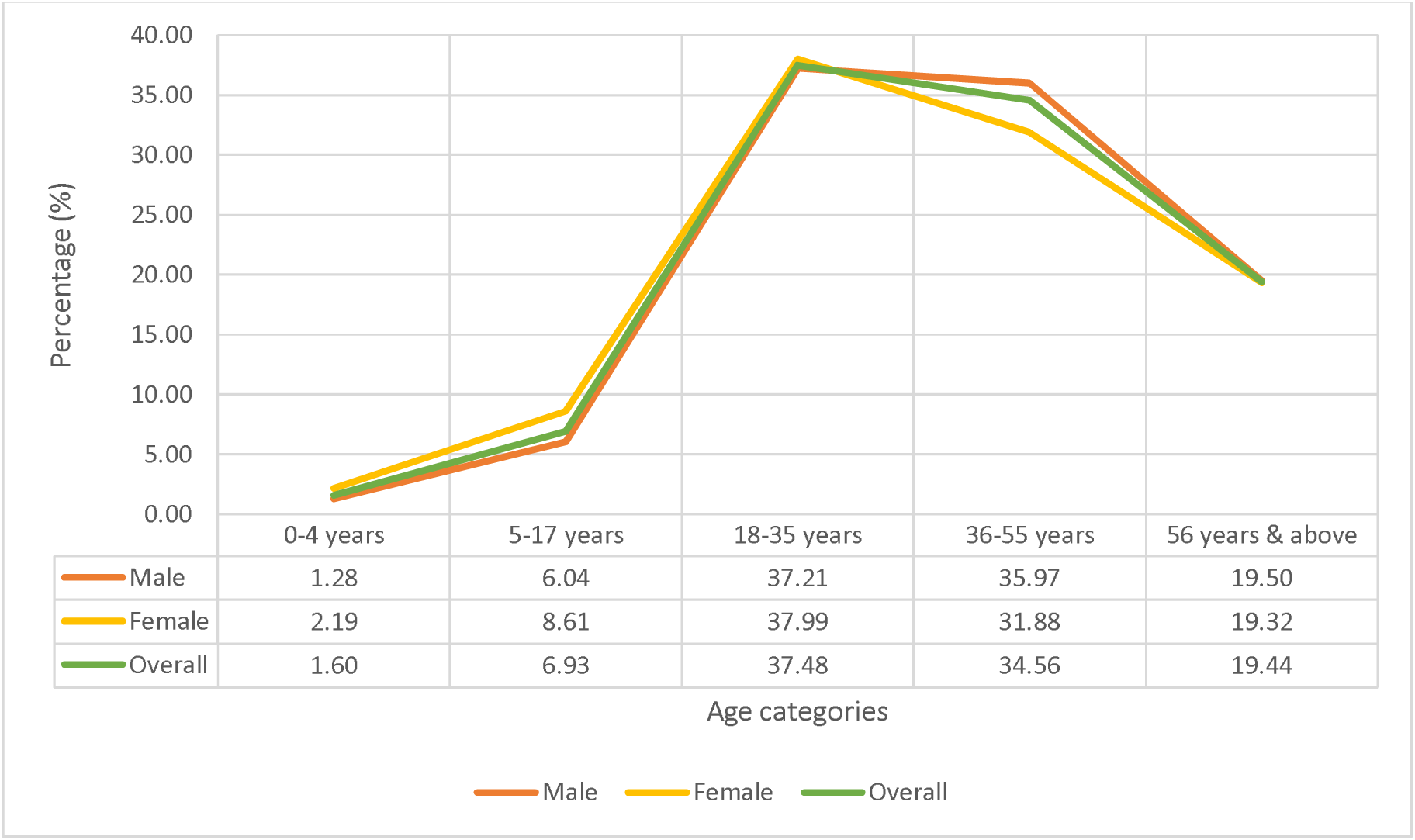
Trends of COVID-19 cases among males and females across different age categories.

The logistic regression analysis between age categories and gender shows significantly higher odds for infection among females in all age categories except for 36-55 years age category after considering 56 years & above age categories as constant. The odds of being COVID-19 infected females was high in 0-4 years (O.R.=1.723, 95% C.I.=1.564-1.898), 5-17 years (O.R.=1.439, 95% C.I.=1.365-1.517) and 18 to 35 years (O.R.=1.030, 95% C.I.=0.996-1.066) age category and it decreases among 36 to 55 years (O.R.=0.894, 95% C.I.=0.864-0.926) age categories (Table 2). Therefore, the odds of COVID-19 infection are higher among females in comparison to males for lower age categories and it decreases with the increasing age.

**Table 2.** Binary logistic regression model for age-categories as the predictor for gender.

| Age Categories | Coefficient | p-value | O.R. | 95% C.I. for O.R. |  |
| --- | --- | --- | --- | --- | --- |
|  |  |  |  | Lower | Upper |
| 0-4 years | 0.544 | <0.000 | 1.723 | 1.564 | 1.898 |
| 5-17 years | 0.364 | <0.000 | 1.439 | 1.365 | 1.517 |
| 18-35 years | 0.030 | 0.087 | 1.030 | 0.996 | 1.066 |
| 36-55 years | -0.112 | <0.000 | 0.894 | 0.864 | 0.926 |
| Constant | -0.645 | <0.000 | 0.525 | ----- |  |

\*Reference category= "56 years& above" \*Predicted probabilities for "females"

Another set of longitudinal analysis was carried out for n=9,131 records which include the patient’s COVID-19 status (recovered/deceased) details along with gender and age. Among the total number of deceased and recovered patients, the proportion of deceased patients increases with age, whereas the proportion of recovered patients increases until 18-35 years (Figure 3). The significant mean age difference (p<0.001) was observed between recovered (36.85±18.51) and deceased (59.99±14.36) categories. Overall patient deceased (94.50%) was greater than the patient recovered (5.50%) in currently assessed records (Supplementary File).

**Figure 3.**
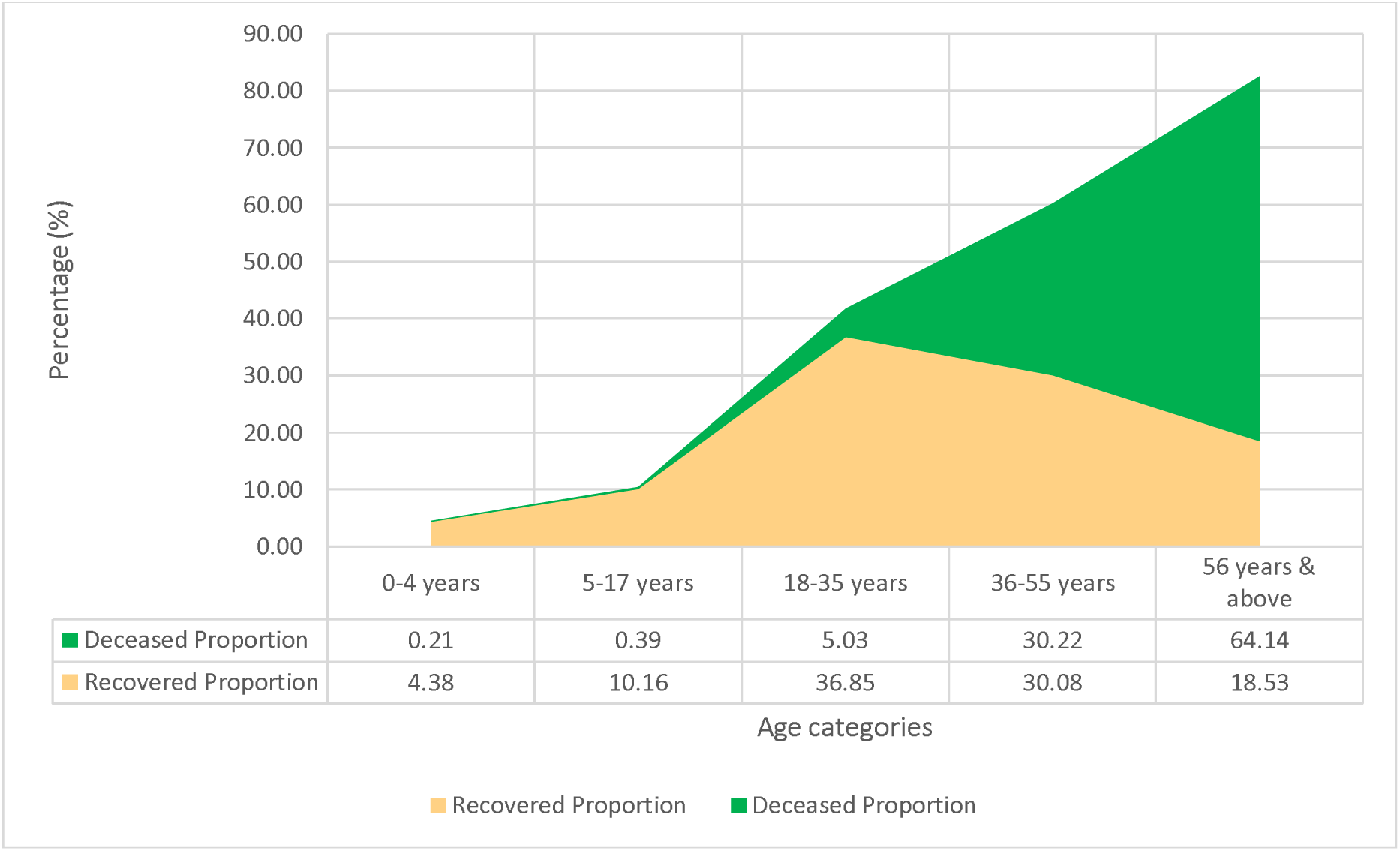
Trends of COVID-19 patient’s status across different age categories.

The binary logistics regression model predicted significant odds of recovery among 0-4 years (O.R.=67.604, 95% C.I.=34.872-131.059), 5-17 years (O.R.=88.286, 95% C.I.=54.423-143.217), 18-35 years (O.R.=25.121, 95% C.I.=19.202-32.865) and 36-55 years (O.R.=3.421, 95% C.I.=2.631-4.450) age categories when 56 years & above age category was considered as constant and adjusted for gender (Table 3). The chances of recovery were higher among females and decreasing age with both adjusted and unadjusted values. The unadjusted results showed ∼8% decrease in odds for being recovered (O.R.=0.919, 95% C.I.=0.914-0.925) with a unit (year) increase in age and about 2 fold higher (O.R.=1.877, 95% C.I.=1.565-2.250) chances of recovery among females (Table 4). However, after adjustment for age, the odds for recovery slightly decreased for females (O.R.=1.765, 95% C.I.=1.442-2.159) and increases for males (O.R.=0.562, 95% C.I.=0.460-0.687).

**Table 3.** Adjusted binary logistic regression model for age and gender categories as predictors for COVID-19 status.

| Categories | Coef. | p-value | O.R. | 95% C.I. for O.R. |  |
| --- | --- | --- | --- | --- | --- |
|  |  |  |  | Lower | Upper |
| Gender | 0.576 | <0.001 | 1.779 | 1.456 | 2.175 |
| 0-4 years | 4.214 | <0.001 | 67.604 | 34.872 | 131.059 |
| 5-17 years | 4.481 | <0.001 | 88.286 | 54.423 | 143.217 |
| 18-35 years | 3.224 | <0.001 | 25.121 | 19.202 | 32.865 |
| 36-55 years | 1.230 | <0.001 | 3.421 | 2.631 | 4.450 |
| Constant | -4.297 | <0.001 | 0.014 | ----- |  |

Reference Category = "males" and "56 years& above"; Predicted probabilities for "recovered".

**Table 4.** Showing adjusted and unadjusted odds of recovery for age (in years) and gender.

|  | Adjusted O.R. (95% C.I.) | Unadjusted O.R. (95% C.I.) |
| --- | --- | --- |
| Age (in years) | 0.920 (0.914 - 0.925) | 0.919 (0.914 - 0.925) |
| Male | 0.562 (0.460 - 0.687) | 0.533 (0.444 - 0.639) |
| Female | 1.765 (1.442 - 2.159) | 1.877 (1.565 - 2.250) |

*\*adjusted for age and gender.*

## Discussion

The SARS-CoV-2 virus being a novel virus can infect human race irrespective of age categories and gender (18). However, there exist individual variations in physiological functions, immune responses and risk factors across gender and age. Therefore, the chances of getting infected might vary among gender and different age categories. The present study attempted to explore the category wise (age and gender) chances of getting infected and predicted chances of getting recovered or deaths with increasing age in the Indian population. In this study, 112,860 COVID-19 patients records were extracted and analysed from a web-based portal to determine the role of age and gender determining COVID-19 status among Indian population. Various studies across the globe show that the older males were more susceptible (>50%) in getting infected by SARS-CoV-2 (16, 19, 20). 60.3% of all cases were found to be males in a study among 5700 hospitalised COVID-19 patients in the U.S. (15). In the present study, the male (65.39%) COVID-19 patients were higher than females (34.61%), and this trend remains consistent among all age categories. The odds ratio shows that the chances for getting recovered among females were high as compared to males independent of age. The researchers consider higher rates of smoking, lower handwashing rates, prior respiratory conditions, biological difference between sexes as a driving force for higher infection and mortality among males (21). The current study suggests higher chances of infection among females with lower age and males with higher age.

Hormonal response elements like putative androgen response elements (AREs) and oestrogen response elements (ORE) produces several innate immunity responses through a genetic mechanism which results in dimorphic innate immunity (22). Several studies emphasized the higher susceptibility of males to viral infection and produce lower antibodies than women. The higher level of TRL7 (Toll-like receptor 7 - protein sensor of RNA viruses) among women produces high interferon-α which provides higher innate immunity to women (23, 24). In other diseases like cancer and HIV, the women also show greater innate immunity and greater response to vaccines (25). The child and adult females show high CD4^+^T cells and CD8^+^T cells and, higher CD4/CD8 ratio and proliferating T-cells compared to males (6, 12). Regardless of age, the females tend to show higher antibody response, immunoglobin levels and B cells which are further enhanced by the genetic factors (presence of an extra X chromosome) (7, 9-11). The regression analysis in the present study depicted age-category wise variation in COIVD-19 cases. The younger females and older males were at higher risk for getting infected and overall chances of recovery decreases as age increases. This may be because the adaptive immunity plays a significant role in response to viral infection and it declines after a certain age which makes us vulnerable to infections. Among males and females, adaptive immunity also varies differentially with age. During the childhood stage or infant stage, the males possess a greater level of IgA, IgM and T_reg_ cells count and an equal number of CD4/CD8 ratio, CD8^+^T cell and B cells. But as the age increases (after puberty/adulthood) the CD4/CD8 ratio, B cells, immunoglobins and T cell proliferation/activation enhances more in females (25). The results can be substantiated and explained based on the evidence that; the male adaptive immune system weakens with increasing age as compared to females.

To extend the current study with additional findings, 9,131 records were separated from previous extracted records based on the patient’s status (recovered/deceased) and analysed. The results of the present study have also significantly predicted the chances of being recovered in different sexes and age categories. The females and 5 to 17 years age category have the highest chances of being recovered followed by 0 to 4 years age category. The prognosis of COVID-19 can be greatly affected by comorbidities and greater risk of developing critical and mortal conditions among male, elderly (>65 years) and smoking patients has been made evident by a study (16), which reflects that both age and gender have an important role in the development of COVID-19 infection. A study among 1590 COVID-19 patients showed a hazard ratio of 1.79 and 2.59 among patients reporting one and two co-morbid conditions (hypertension and diabetes), respectively (26). NK (natural killer) cells increase greatly among females with age as compared to males and they experience a decline in lymphocytes elements during/after menopause, but the elderly males experience a greater rapid decline in B cell count, CD4^+^T cells and T cell proliferation than females (11, 27). In recent studies, males were reported to have increased level of plasma ACE2 concentration (28) and ACE2 is the receptor required for cellular entry of SARS CoV-2 (29). The plasma ACE2 level was found to be highly correlated with immune signatures in lungs of males and older person and less correlated among females and younger person (29). So, the chances of infection and death due to COVID-19 was higher among elderly males. This also has been corroborated by the results of the present study, where the chances of being COVID-19 infected were observed to be higher for males with increasing age and chances of recovery increases with decrease in age and for females. After adjustment for age, the chances of recovery from COVID-19 among females slightly decreases whereas it increases for males. Another perspective for higher infection among males of higher ages was X and Y chromosomes-based variations. Both these chromosomes harbours gene which involved in secreting the immune response elements but in females mosaic form of X chromosomes results into heterogenic ACE2 allele. Thus, efficient form of ACE2 receptor is present only in half of all cells which limit the infection/attachment of SARS-CoV-2 virus and provides relatively greater protection to females (30).

## Conclusion

The chances of getting infected with SARS-CoV-2 varies with age-categories and gender. The females of lower age categories (<35 years) have higher chances of getting infected than male counterparts and as age increases (>35 years) the infection rate for male increases. The chances of getting recovered declines with increasing age and it is lower among males as compared to females. The variation in infection rates and chances of recovery and deaths across age and gender might be based on the biological variations like immunological and genetic differences, which however needs further verifications, considering both social and biological factor separately. The public health policies and new treatment strategies can be developed considering the present findings.

## Data Availability

The data used in this manuscript is available via Mendeley data

http://dx.doi.org/10.17632/d6x59tgss8.1

## Strength and Limitations

The present study provides preliminary estimates of the association of age and gender with COVID-19 cases in India which were rarely available. Evidence to correlate the role of the various immunological and genetic basis of the observed variations is offered by the present study for pursuing research. The data used for the current study was from a private source, nevertheless, these data were also used by a government institute for the development of COVID-19 prediction portal (http://covid-tracker.iimv.ac.in:3939/covid/) therefore, the validity of using the aforementioned data persists.

## Acknowledgements

We would like to acknowledge all the associated persons involved in updating https://api.covid19india.org/documentation/csv/ web portal.

## Ethical Approval

The authors have used secondary data source for developing the present article therefore, ethical approval was not required.

## Funding Source

None

## Notes

### Competing Interest Statement

The authors have declared no competing interest.

### Funding Statement

No funding source

### Author Declarations

The IRB body of Department of Community Medicine and School of Public Health, PGIMER, Chandigarh, India approved this research.

## References

1. WHO Coronavirus Disease (COVID-19) Dashboard 2020. Available from: https://covid19.who.int/?gclid=Cj0KCQjww_f2BRC-ARIsAP3zarH9e-pD1WIUN8gqV1mMbbv8UtsUDSv0CdxxhklRSF1u7XZNzJsVg4YaAsUyEALw_wcB.

2. Sarkar A, Chouhan P. COVID-19: District level vulnerability assessment in India. Clinical Epidemiology and Global Health. 2020. doi:10.1016/j.cegh.2020.08.017.

3. Lulbadda KT, Kobbekaduwa D, Guruge ML. The impact of temperature, population size and median age on COVID-19 (SARS-CoV-2) outbreak. Clinical Epidemiology and Global Health. 2020. doi:10.1016/j.cegh.2020.09.004.

4. Wasdani KP, Prasad A. The impossibility of social distancing among the urban poor: the case of an Indian slum in the times of COVID-19. Local Environment. 2020;25(5):414–8. doi:10.1080/13549839.2020.1754375.

5. Sengupta S, Jha MK. Social Policy, COVID-19 and Impoverished Migrants: Challenges and Prospects in Locked Down India. The International Journal of Community and Social Development. 2020;2(2):152–72. doi:10.1177/2516602620933715.

6. Lee BW, Yap HK, Chew FT, Quah TC, Prabhakaran K, Chan GSH, et al. Age-and Sex-Related changes in lymphocyte subpopulations of healthy Asian subjects: From birth to adulthood. Communications in Clinical Cytometry. 1996;26:8–15. doi:10.1002/(SICI)1097-0320(19960315)26:1<8::AID-CYTO2>3.0.CO;2-E. PubMed PMID: 8809475.

7. Furman D, Hejblum BP, Simon N, Jojic V, Dekker CL, Thiebaut R, et al. Systems analysis of sex differences reveals an immunosuppressive role for testosterone in the response to influenza vaccination. Proceedings of the National Academy of Sciences of the United States of America. 2014;111:869–74. doi:10.1073/pnas.1321060111. PubMed PMID: 24367114.

8. Simon AK, Hollander GA, McMichael A. Evolution of the immune system in humans from infancy to old age. Proceedings of the Royal Society B: Biological Sciences. 2015;282(1821):20143085. doi:10.1098/rspb.2014.3085.

9. Abdullah M, Chai PS, Chong MY, Tohit ERM, Ramasamy R, Pei CP, et al. Gender effect on in vitro lymphocyte subset levels of healthy individuals. Cellular Immunology. 2012;272:214–9. doi:10.1016/j.cellimm.2011.10.009. PubMed PMID: 22078320.

10. Fan H, Dong G, Zhao G, Liu F, Yao G, Zhu Y, et al. Gender differences of B cell signature in healthy subjects underlie disparities in incidence and course of SLE related to estrogen. Journal of Immunology Research. 2014;2014:1–17. doi:10.1155/2014/814598. PubMed PMID: 24741625.

11. Teixeira D, Longo-Maugeri IM, Santos JLF, Duarte YAO, Lebrão ML, Bueno V. Evaluation of lymphocyte levels in a random sample of 218 elderly individuals from São Paulo city. Revista Brasileira de Hematologia e Hemoterapia. 2011;33:367–71. doi:10.5581/1516-8484.20110100.

12. Uppal SS, Verma S, Dhot PS. Normal Values of CD4 and CD8 Lymphocyte Subsets in Healthy Indian Adults and the Effects of Sex, Age, Ethnicity, and Smoking. Cytometry Part B - Clinical Cytometry. 2003;52:32–6. doi:10.1002/cyto.b.10011. PubMed PMID: 12599179.

13. Zhou F, Yu T, Du R, Fan G, Liu Y, Liu Z, et al. Clinical course and risk factors for mortality of adult inpatients with COVID-19 in Wuhan, China: a retrospective cohort study. The Lancet. 2020;395:1054–62. doi:10.1016/S0140-6736(20)30566-3. PubMed PMID: 32171076.

14. Zhang H, Penninger JM, Li Y, Zhong N, Slutsky AS. Angiotensin-converting enzyme 2 (ACE2) as a SARS-CoV-2 receptor: molecular mechanisms and potential therapeutic target. Intensive Care Medicine. 2020;46:586–90. doi:10.1007/s00134-020-05985-9. PubMed PMID: 32125455.

15. Richardson S, Hirsch JS, Narasimhan M, Crawford JM, McGinn T, Davidson KW, et al. Presenting Characteristics, Comorbidities, and Outcomes Among 5700 Patients Hospitalized With COVID-19 in the New York City Area. JAMA. 2020;323:2052. doi:10.1001/jama.2020.6775.

16. Jin J-MM, Bai P, He W, Wu F, Liu X-FF, Han D-MM, et al. Gender Differences in Patients With COVID-19: Focus on Severity and Mortality. Frontiers in Public Health. 2020;8:152. doi:10.3389/fpubh.2020.00152.

17. Kushwaha S. COVID-19_AGE_GENDER_STATUS_INDIA_Dataset 2020.

18. Zhonghua CN-Zlxbxz. The epidemiological characteristics of an outbreak of 2019 novel coronavirus diseases (COVID-19) in China. cmajissn. 2020. doi:10.3760/cma.j.issn.0254-6450.2020.02.003.

19. Mallapaty S. The coronavirus is most deadly if you are older and male - new data reveal the risks. Nature. 2020;585:16–7. doi:10.1038/d41586-020-02483-2. PubMed PMID: 32860026.

20. Munayco C, Chowell G, Tariq A, Undurraga EA, Mizumoto K. Risk of death by age and gender from CoVID-19 in Peru, March-May, 2020. Aging. 2020;12:13869–81. doi:10.18632/aging.103687. PubMed PMID: 32692724.

21. Betron M, Gottert A, Pulerwitz J, Shattuck D, Stevanovic-Fenn N. Men and COVID-19: Adding a gender lens. Global Public Health. 2020;15:1090–2. doi:10.1080/17441692.2020.1769702. PubMed PMID: 32436422.

22. Hannah MF, Bajic VB, Klein SL. Sex differences in the recognition of and innate antiviral responses to Seoul virus in Norway rats. Brain, Behavior, and Immunity. 2008;22:503–16. doi:10.1016/j.bbi.2007.10.005. PubMed PMID: 18053684.

23. Berghöfer B, Frommer T, Haley G, Fink L, Bein G, Hackstein H. TLR7 Ligands Induce Higher IFN-α Production in Females. The Journal of Immunology. 2006;177:2088–96. doi:10.4049/jimmunol.177.4.2088. PubMed PMID: 16887967.

24. Pisitkun P, Deane JA, Difilippantonio MJ, Tarasenko T, Satterthwaite AB, Bolland S. Autoreactive B cell responses to RNA-related antigens due to TLR7 gene duplication. Science. 2006;312:1669–72. doi:10.1126/science.1124978. PubMed PMID: 16709748.

25. Klein SL, Flanagan KL. Sex differences in immune responses. Nature Reviews Immunology. 2016;16:626–38. doi:10.1038/nri.2016.90. PubMed PMID: 27546235.

26. Nelson CP, Sama IE, Codd V, Webb TR, Ye S, Lang CC, et al. Genetic Associations With Plasma Angiotensin Converting Enzyme 2 Concentration: Potential Relevance to COVID-19 Risk. Circulation. 2020;142:1117–9. doi:10.1161/CIRCULATIONAHA.120.049007. PubMed PMID: 32795093.

27. Hirokawa K, Utsuyama M, Hayashi Y, Kitagawa M, Makinodan T, Fulop T. Slower immune system aging in women versus men in the Japanese population. Immunity & Ageing. 2013;10. doi:10.1186/1742-4933-10-19.

28. Osama T, Pankhania B, Majeed A. Protecting older people from COVID-19: should the United Kingdom start at age 60? Journal of the Royal Society of Medicine. 2020. doi:10.1177/0141076820921107.

29. Li MY, Li L, Zhang Y, Wang XS. Expression of the SARS-CoV-2 cell receptor gene ACE2 in a wide variety of human tissues. Infectious Diseases of Poverty. 2020;9:45. doi:10.1186/s40249-020-00662-x. PubMed PMID: 32345362.

30. Gemmati D, Bramanti B, Serino ML, Secchiero P, Zauli G, Tisato V. COVID-19 and individual genetic susceptibility/receptivity: Role of ACE1/ACE2 genes, immunity, inflammation and coagulation. might the double x-chromosome in females be protective against SARS-COV-2 compared to the single x-chromosome in males? International Journal of Molecular Sciences. 2020;21:3474. doi:10.3390/ijms21103474. PubMed PMID: 32423094.

