## Supplementary File for "Biological attributes of age and gender variations in Indian COVID-19 cases: A retrospective data analysis"

Supplementary File Table

Table. Showing the mean age difference in the patient`s COVID-19 status categories.

| Patient`s Status | n (%) | Mean ± S.D. | T-test | p-value |
| --- | --- | --- | --- | --- |
| Recovered | 502 (5.50) | 36.85±18.51 | 34.48 | <0.001 |
| Deceased | 8629 (94.50) | 59.99±14.36 |  |  |
| Overall | 9131 (100.00) | 58.72±15.54 | --------------------- | |
